# Unexpected Transmission Dynamics in a University Town: Lessons from COVID-19

**DOI:** 10.1101/2024.01.10.24301116

**Authors:** Erin Clancey, Matthew S. Mietchen, Corrin McMichael, Eric T. Lofgren

## Abstract

Institutions of higher education faced a number of challenges during the COVID-19 pandemic. Chief among them was whether or not to re-open during the second wave of COVID-19 in the fall of 2020, which was controversial because incidence in young adults was on the rise. The migration of students back to campuses worried many that transmission within student populations would spread into surrounding communities. In light of this, many colleges and universities implemented mitigation strategies, with varied degrees of success. Washington State University (WSU), located in the city of Pullman in Whitman County, WA, is an example of this type of university-community co-location, where the role of students returning to the area for the fall 2020 semester was contentious. Using COVID-19 incidence reported to Whitman County, we retrospectively study the transmission dynamics that occurred between the student and community subpopulations in fall 2020. We develop a two-population ordinary differential equation mechanistic model to infer transmission rates within and across the university student and community subpopulations. We use results from Bayesian parameter estimation to determine if sustained transmission of COVID-19 occurred in Whitman County and the magnitude of cross-transmission from students to community members. We find these results are consistent with estimation of the time-varying reproductive number and conclude that the students returning to WSU-Pullman did not place the surrounding community at disproportionate risk of COVID-19 during fall 2020 when mitigation efforts were in place.

## Introduction

The COVID-19 pandemic dramatically reshaped pedagogy in higher education [1, 2, 3], impacted the mental health and behavior of university students [4], restructured institutional response and governance within colleges and universities [1, 3], and forced difficult decisions on students and their families [5]. Looking back, decisions affecting both public health and education were difficult to balance [6, 7]. University (or college) towns, communities that are socioeconomically dominated by a college or university, pose unique public health challenges because they house populations with different demographics and interests that are regulated under common policy. Campus closures and online learning that potentially reduced transmission of acute respiratory syndrome coronavirus 2 (SARS-CoV-2), arguably had an adverse affect on many college students [8]. At the same time, universities were thought to have intensified transmission [9, 10], with high incidence among student populations conceivably putting non-student community residents at greater risk [11, 12]. During the pandemic, colleges and universities were labeled as hot spots of virus transmission [13] and the re-opening of college campuses, particularly for the fall 2020 semester, was controversial [11, 6] with different institutions adopting a wide variety of mitigation and teaching strategies [12, 14]. Even in the aftermath, the precise role that college-age and university students played transmission amongst the greater community remains up for debate [see 8].

Locally, college and university student populations were pivotal in the spread of SARS-CoV-2 when campuses re-opened in the fall of 2020 for two overarching reasons, mobility and social mixing [8]. By August 2020, incidence across the United States was in decline except for young adults, whose demographic saw an increase in incidence with outbreaks reported at many colleges and universities [12, 14, 10]. As campuses re-opened for the fall 2020 semester, many students migrated back to university towns and resumed communal living. Both factors, mobility and social mixing, contributed to increasing incidence by concentrating imported cases on college campuses with the potential for onward transmission in residential and social settings directly linked or adjacent to life on campus [11]. Even though increased incidence put the health of students, faculty and staff, and the surrounding community at greater risk of infection with SARS-CoV-2, it was not guaranteed that increased incidence in students translated directly to increased transmission from students to community residents.

In anticipation of re-opening campuses in the face of high COVID-19 incidence, most colleges and universities increased mitigation and testing efforts to offset the potential for increased transmission [12] and were successful. For example, the University of Illinois at Urbana-Champaign implemented the multi-modal ‘SHIELD: Target, Test, and Tell’ program along with other non-pharmaceutical interventions in fall 2020 and found these mitigation strategies reduced transmission, hospitalizations and deaths [15]. Several other studies focusing on transmission between student and co-located resident populations demonstrated, using either mobility data [e.g., 14] or genomic surveillance data [e.g., 9, 16, 17], that cross-transmission was highly limited. In contrast, other findings still argue campus outbreaks translated into peaks of infection within their home communities [e.g., 6]. Thus, it is important to retrospectively study transmission dynamics of SARS-CoV-2 in university towns to optimize mitigation strategies for the future.

Here we use mathematical modeling and Bayesian parameter estimation to study transmission dynamics of SARS-CoV-2 within a university student population and a co-located resident community population. Specifically, we study the outbreak of COVID-19 that occurred at Washington State University (WSU) during fall 2020. We develop a two-population compartmental model to estimate transmission that occurred within the student and community populations and cross-transmission between populations using COVID-19 incidence reported to Whitman County, Washington. We also estimate the time-varying reproductive number to make a comparison of a real-time estimation to our retrospective transmission estimates. With the posterior distributions generated from Bayesian model fitting, we address two questions motivated by uncertainty around the potential risk university student populations pose to surrounding communities during epidemics of respiratory disease: (1) Was the COVID-19 outbreak that occurred during fall 2020 in Whitman County a result of sustained transmission within and across the university student and community subpopulations? (2) If cross-transmission did occur, what was the magnitude of population mixing?

## Methods

### Study population and COVID-19 case reporting

Whitman County, located in a rural agricultural area of southeastern Washington, is home to WSU and the city of Pullman. WSU is a large, public research land-grant university, and draws its student body from all over Washington State and beyond. The city of Pullman is a quintessential university town and the largest city in Whitman County. Although the Whitman County community residents and WSU student populations overlap geographically, the two subpopulations do not mix randomly. Students are largely concentrated in housing on or near the WSU campus — ‘College Hill’, while community members live in other areas of the city of Pullman or are dispersed throughout Whitman County.

Far from the Seattle area metropolis, Whitman County was less affected than western Washington by the pandemic in early 2020, but lockdown measures and masking requirements were implemented at both WSU and Pullman city public spaces and many private businesses as part of statewide actions. All WSU courses were still fully remote in fall 2020 and the campus was officially closed for student housing save for special exemptions, however, many students returned to the area, living in apartments or Greek housing near the campus. During this time Whitman County experienced a sharp, dramatic rise in reported COVID-19 cases, primarily among those associated with WSU. Within the first three weeks of the WSU fall 2020 semester, Whitman County reported an outbreak of COVID-19 within the student and subsequently the community populations with one of the highest rates reported in Washington State and nationally at the time [18]. COVID-19 testing was available for both community residents and WSU students throughout the outbreak. University testing sites, in collaboration with local public health, enabled all returning students, faculty, and staff to receive free testing before and after the beginning of the semester. While some negative test requirements were in place, many of these relied on self-attestation and arrival testing was not mandated for students. For Whitman County citizens, testing was available throughout the city for residents at clinics, pharmacies, and non-permanent testing sites. Positive COVID-19 cases were reported to Whitman County public health. The epidemic weekly time series data we present here begins on August 17, 2020, the week before the first day of the WSU fall semester, and reports until December 27, 2020 (the time series is congruent with weeks 34-52). Case reports for each week include the totals for the student and community subpopulations.

### Mathematical model and ℛ_0_

To study the transmission dynamics that occurred in Whitman County during the COVID-19 outbreak of fall 2020, we developed a metapopulation model with explicit intra- and inter-community interactions. Specifically, we used a two-population Susceptible-Exposed-Infected-Recovered (SEIR) ordinary differential equations (ODE) model framework with transmission occurring within the university student, *u*, and Whitman County community, *c*, subpopulations and cross-transmission between these two subpopulations. We assumed the latency rate (*σ*) and recovery rate (*γ*) were equivalent in both populations, and cross-transmission (*β*_*m*_) from university students to community member was equal to transmission from community members to university students. We also assumed that population sizes, *N*, remained constant throughout the duration of the 2020 fall semester, with no substantial loss to death or migration once the students arrived back on campus. Figure (1) is a diagram of the mechanistic model, definitions of model parameters and their symbols are given in Table 1, and the ODE’s specifying the model are given in Appendix A.1 of the Supplement.

**Table 1:** Model symbols for subscripts, variables, parameters and their definitions. Values are given for initial starting conditions for each variable or for fixed parameter values with references. All rates are in weeks unless specified otherwise.

| Symbol | Definition | Initial or Fixed Value | Reference |
| --- | --- | --- | --- |
| $u, c$ | University student or Community member | NA | |
| $\beta_u$ | Transmission rate between university students | Estimated | |
| $\beta_c$ | Transmission rate between community members | Estimated | |
| $\beta_m$ | Cross-transmission rate between subpopulations | Estimated | |
| $\sigma$ | Latency rate | 7/3.59 | [26] |
| $\gamma$ | Rate of recovery from infection | 7/3.56 | [26] |
| $\rho$ | Positive testing (ascertainment) rate | 0.76 | [25] |
| $k$ | Overdispersion parameter | Estimated | |
| $N_u, N_c$ | Subpopulation sizes | 14,254, 20,785 | |
| $S_{u_0}, S_{c_0}$ | Initial number of susceptibles | $N_{u,c} - E_{u_0,c_0} - I_{u_0,c_0} - R_{u_0,c_0}$ | |
| $E_{u_0}, E_{c_0}$ | Initial number of exposed | $\text{Case Reports}_{u_0,c_0} \times \rho^{-1}$ | [25] |
| $I_{u_0}, I_{c_0}$ | Initial number of infected | $\text{Case Reports}_{u_0,c_0} \times \rho^{-1}$ | [25] |
| $R_{u_0}, R_{c_0}$ | Initial number of recovered | $N_{u,c} \times 0.041$ | [25] |

**Figure 1:**
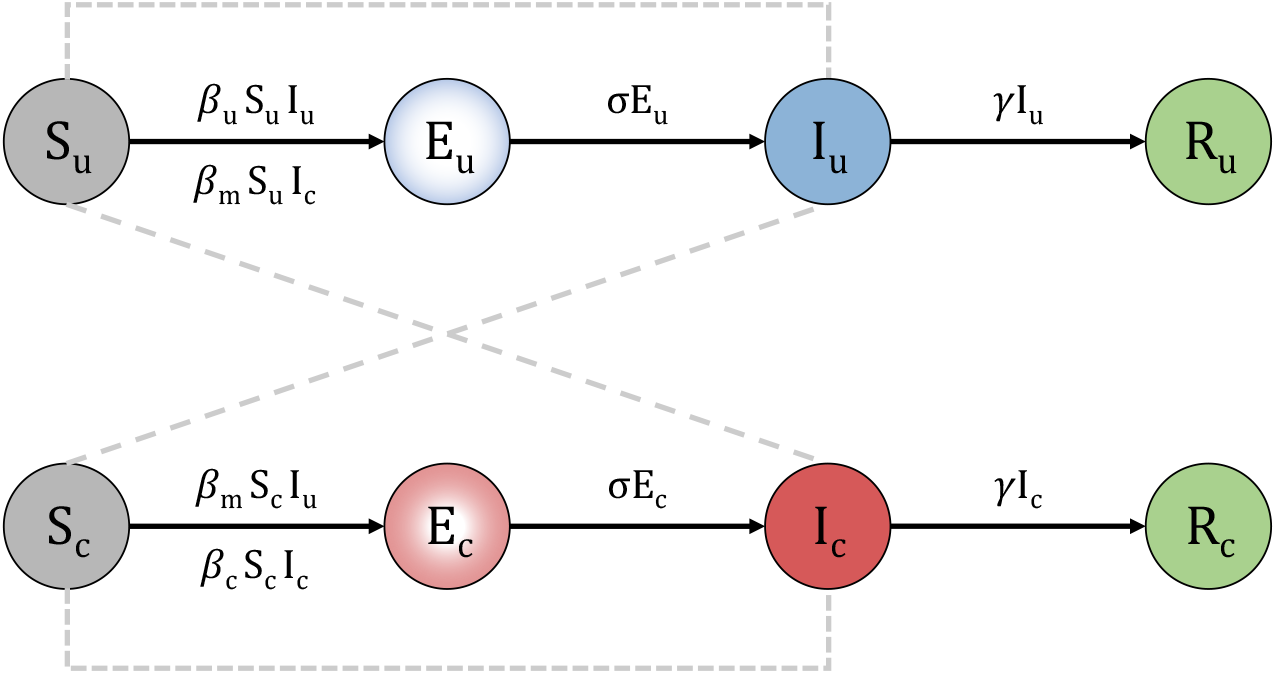
Diagram of the two-population Susceptible-Exposed-Infected-Recovered (SEIR) model. The subscript *u* specifies the university student population and the subscript *c* specifies the Whitman County community population. Parameters represented include the within subpopulation transmission rates, *β*_*u*_ and *β*_*c*_, the cross-transmission rate, *β*_*m*_, the latency rate, *σ*, and the recovery rate *γ*.

The basic reproductive number, ℛ_0_, is a key epidemic parameter that quantifies the transmissibility of a pathogen in a completely susceptible population. We use ℛ_0_ here to determine the potential for sustained transmission in each subpopulation and the total population. We quantify the reproductive numbers for each subpopulation, 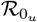 or 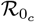, considering only local transmission such that 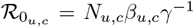, where *N* is the total subpopulation size, and therefore initial size of susceptibles, in either *u* or *c*. We derived 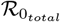 using the next-generation matrix (NGM) following [19] from our two-population SEIR model. Details of the derivation are given in Appendix A.2 of the Supplement. The resulting equation is

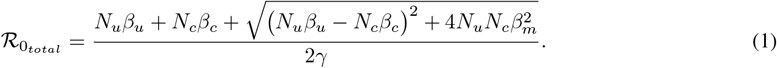

### ℛ_*t*_ Estimation with EpiEstim

Very similar to ℛ_0_, the time-varying reproductive number, ℛ_*t*_, is the average number of secondary infections generated by a single individual over the infectious period at time *t* [20]. Monitoring the reproductive number as an outbreak progresses can provide instantaneous feedback to public health officials on the effectiveness of control measures [20]. The `EpiEstim` framework developed by Cori et al. [20] is a popular method to estimate ℛ_*t*_ from case data and is freely available as an `R` package [21, 22]. `EpiEstim` is simple to implement, requiring only incidence time series and a generation (or serial) interval distribution. In addition to daily incidence data, `EpiEstim` can utilize temporally aggregated data even when the time window of incidence reporting is longer than the mean generation interval (e.g., when incidence is reported over weekly intervals or aggregated to reduce administrative noise, such as the effects of the weekends) [23]. Even when incidence data is temporally aggregated, the resulting ℛ_*t*_ estimates are made daily. Here we use `EpiEstim` to estimate ℛ_*t*_ directly from the Whitman County weekly aggregated case reports for the total population and for each subpopulation. We use the generation interval distribution derived directly from our two-population ODE model (see Appendix A.3 of the Supplement) with the values for *σ* and *γ* from Table 1. These data-driven estimates provide a useful comparison to our model-based estimates of ℛ_0_ to understand if sustained transmission occurred in Whitman County during fall 2020.

### Statistical model and Bayesian inference

To understand the COVID-19 transmission dynamics in Whitman County, we used Bayesian approaches to estimate the three transmission parameters, *β*_*u*_, *β*_*c*_, and *β*_*m*_ from our two-population model. We implemented the simulation-based estimation methods available in the `pomp` library in R [24, 22] to generate maximum *a posteriori* (MAP) estimates and 95% highest posterior density interval (HPD) intervals. The `pomp` framework required that we also specify a statistical distribution to model the sampling process that generated the case reporting data in addition to our two-population model. For this we used a negative binomial model, which includes two additional parameters, *ρ*, the positive testing rate, and *k*, the overdispersion parameter (see Appendix B.1 of the Supplement for details on the negative binomial likelihood function). We fixed *ρ* using the ascertainment rate estimated for Washington State during the first two weeks of September 2020 from [25] (Table 1), but estimated *k*. Initializing the simulation-based methods also required specifying values for *S, E, I*, and *R* for the university student and community populations at time zero. To approximate the initial values 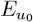, 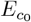, 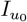, and 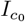 we divided the number of reported COVID-19 cases from week 34 (the week before classes began in fall 2020 and when students usually arrive back in Whitman County) by *ρ*. To approximate the initial values for 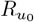 and 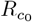 we multiplied the the subpopulation sizes (*N*_*u*_, *N*_*c*_) by the estimated number of total COVID-19 cases as of September 3, 2020 in Washington State from [25] (Table 1).

## Results

We begin our analyses of the Whitman County fall 2020 COVID-19 outbreak by focusing on the observed case reports. Figure (2) shows the weekly cases reported to Whitman County beginning on week 34 of the calendar year (the week before classes commenced at WSU) for the total population and for each subpopulation. Figure (2) also shows the ℛ_*t*_ estimates from `EpiEstim` for the total population and each subpopulation. ℛ_*t*_ estimated for the total population begins just over 3, falls to 1 by week 252 (week 37), and then hovers near 1 throughout the duration of the fall semester. The subpopulations demonstrate more fluctuations, however, ℛ_*t*_ estimated for the university student population is similar to the total population. ℛ_*t*_ estimated for the WSU students also begins near 3, falls below 1 by day 252 (week 37), and remains near or below one until the end of the semester with a spike to near 3 on day 327 (week 47, the week before Thanksgiving break). ℛ_*t*_ estimated for the community subpopulation does not begin until day 279 (week 41) because the observed case counts are below 11 reports per week and estimates from `EpiEstim` are unreliable when cases drop below 11 [20]. ℛ_*t*_ estimated for the community subpopulation begins just above 4 and falls to one by day 297 (week 43). Then, a spike similar to the WSU estimates occurs in in the community with a peak on day 320 (the last day of week 46), and then returns to near 1 for the remainder of the semester.

**Figure 2:**
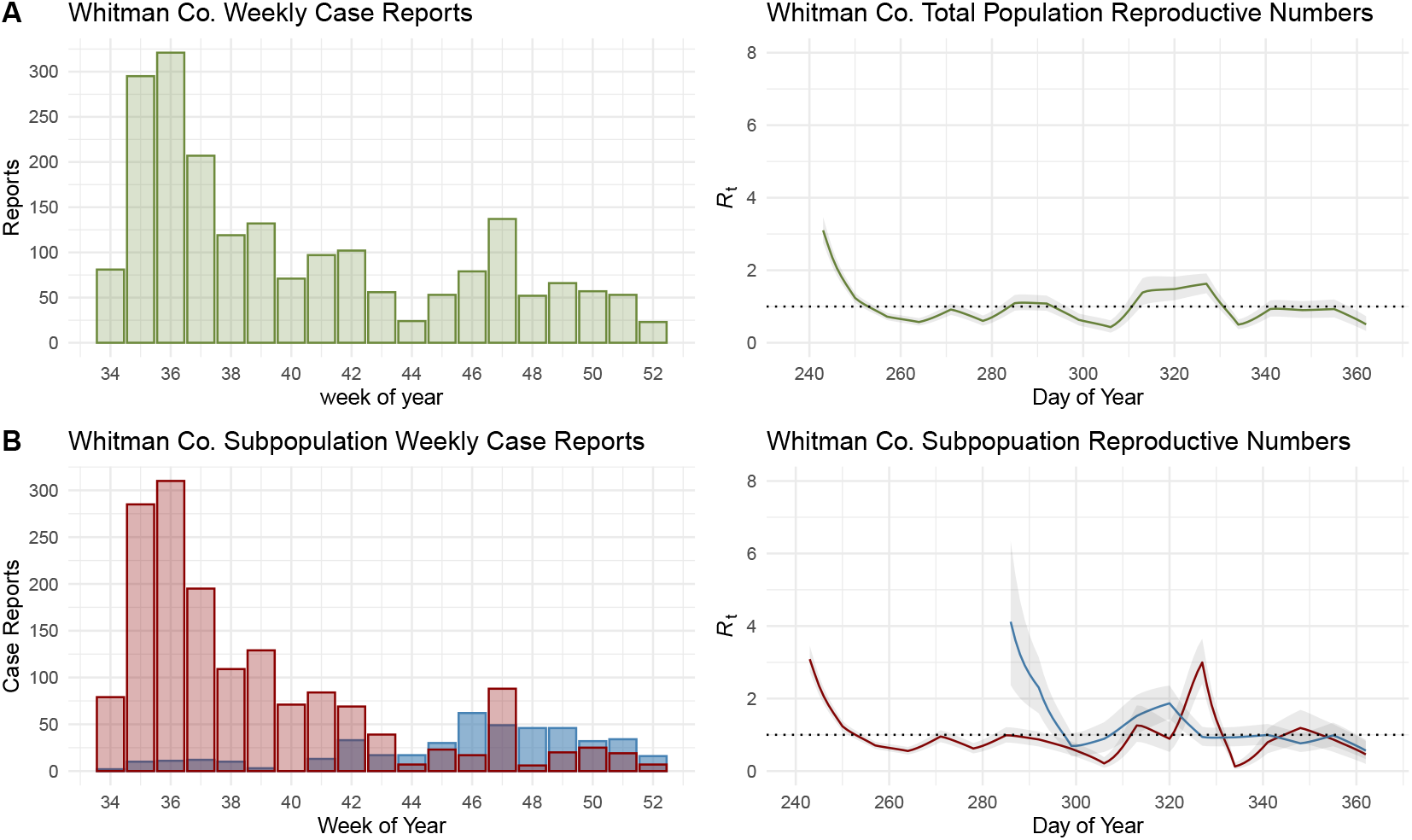
Whitman Co. fall 2020 COVID-19 weekly case reports and ℛ_*t*_ daily estimates from the total population shown in the top row (panel **A**) and the university student (red) and Whitman Co. community (blue) subpopulations shown in the bottom row (panel **B**). Horizontal dotted lines cross the y-axis at 1 for ℛ_*t*_ daily estimates.

Next, using Bayesian inference to estimate the three transmission parameters (*β*_*u*_, *β*_*c*_, *β*_*m*_) and ℛ_0_ in the total population and both subpopulations, we investigate if sustained transmission occurred within each subpopulation and quantify the magnitude of cross-transmission across subpopulations. Transmission was significantly greater in the university student population than in the community (Figure 3), as the 95% HPD intervals are not overlapping (Table2). Cross-transmission was much less than local transmission within either subpopulation, but was significantly greater than zero (Table 2). All 95% HPD intervals for local and total population ℛ_0_’s overlap, however, the posterior distribution for 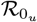 does include the critical threshold of one. Nevertheless, since all ℛ_0_’s are near or below one, sustained transmission of COVID-19 was likely very weak during fall 2020 in Whitman County. The overdispersion parameter *k* was unknown and therefore estimated, but it not the focus of this study. Values for this parameter are reported in Table (2) and the posterior distribution is shown in Appendix B.2 in the Supplement.

**Table 2:** Results from Bayesian estimation with pomp for each estimated parameter from Table 1 and ℛ_0_. Maximum a posteriori (MAP) estimates are modes of the marginal posterior distributions with 95% highest posterior density (HPD) credible intervals.

| Parameter | MAP estimate | 95% HPD intervals |
| --- | --- | --- |
| $\beta_u$ | $1.23 \times 10^{-4}$ | $[1.12 \times 10^{-4}, 1.32 \times 10^{-4}]$ |
| $\beta_c$ | $8.95 \times 10^{-5}$ | $[7.59 \times 10^{-5}, 1.05 \times 10^{-4}]$ |
| $\beta_m$ | $1.69 \times 10^{-6}$ | $[0.00, 4.00 \times 10^{-6}]$ |
| $k$ | 2.43 | $[1.29, 4.51]$ |
| $\mathcal{R}_{0_u}$ | 0.90 | $[0.81, 0.95]$ |
| $\mathcal{R}_{0_c}$ | 0.95 | $[0.80, 1.11]$ |
| $\mathcal{R}_{0_{total}}$ | 0.94 | $[0.86, 1.10]$ |

**Figure 3:**
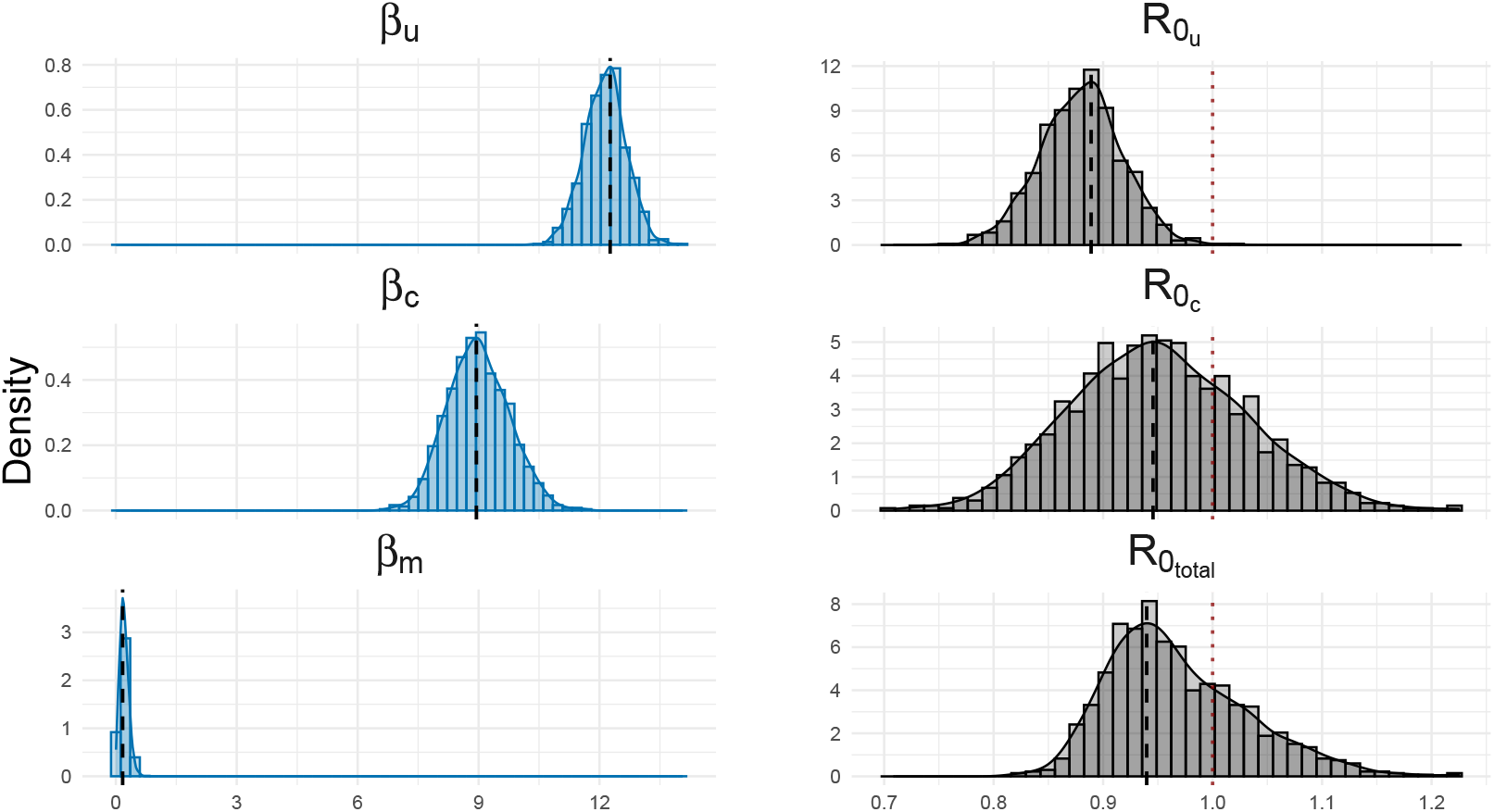
Posterior densities for each transmission parameter and ℛ_0_ for the university student and community subpopulations and the total population. Vertical black dashed lines represent the maximum a posteriori (MAP) estimate. Vertical red dotted lines delineate the critical threshold where ℛ_0_ = 1.

## Discussion

Our retrospective analyses investigated the transmission dynamics of SARS-CoV-2 that occurred between WSU students and the surrounding community during fall 2020. A major strength of our analyses stems from the of pairing COVID-19 incidence data with a mechanistic model. This approach allowed for direct estimation of transmission and cross-transmission rates within and across the university student and community subpopulations, and for the estimation of ℛ_0_ for the university student, community and total populations. We find that sustained global or local transmission of SARS-CoV-2 infection did not occur in Whitman County in fall 2020 even in the face of student movement back to the WSU-Pullman campus. The magnitude of cross-transmission between the university student and community subpopulations was small and not significantly different from zero. Our results demonstrate support for non-pharmaceutical interventions and social distancing policies effectively reducing transmission of SARS-CoV-2 within each subpopulation, and the inherent non-random mixing between university students and surrounding communities further reducing transmission between the geographically co-located subpopulations.

In late August 2020, many university students returned to the WSU campus in Pullman, Washington even though course delivery was completely remote. Arrival testing was not mandatory for returning students and therefore identifying imported cases was not possible. However, importation of COVID-19 cases likely occurred with student immigration back to Whitman County. Student COVID-19 incidence peaked in the second week of the fall semester and then immediately began falling. This is in contrast, for example, to the outbreak at an Arkansas university reported in [10], which demonstrated rising cases after classes started on August 24, 2020 and not falling until early September after an Arkansas department of health testing event. Similarly at the University of Michigan–Ann Arbor, COVID-19 incidence increased throughout the fall 2020 semester and peaked in mid-November [16]. Even though the COVID-19 epidemic at the University of Michigan–Ann Arbor was a result of sustained transmission among the student population, Valesano et al. [16] concluded that the outbreak was derived from multiple imported cases with very little spread into the community. Transmission dynamics at WSU during fall 2020 had noticeably different characteristics compared to these other universities that experienced outbreaks with onward transmission. Some transmission did occur at WSU occur, which was detected in the ℛ_*t*_ estimates. However, 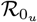 was estimated to be significantly less than 1, and ℛ_*t*_ estimates for the WSU students dropped to 1 quickly and remained near 1 for the duration of the fall semester. However, like other universities, importation could have still been in effect and transmission from WSU students to community members was minimal.

Even with the return of WSU students and increased COVID-19 incidence, the Whitman County community did not experience a measurable heightened risk of COVID-19 in fall 2020. Our results demonstrate limited cross-transmission between university students and the surrounding community. There was a distinctive delay in the progression of COVID-19 cases in the community after the beginning of the fall 2020 semester with the peak in student incidence occurring asynchronously with peak incidence in the community. Case numbers in the community did not begin to rise until late-October and early-November with ℛ_*t*_ estimation not being possible until week 41. Overall, transmission within the community subpopulation was minimal, with 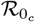 near 1, and the interaction with the university student supopulation did not increase 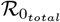 to be statistically greater than 1, which is consistent with the ℛ_*t*_ from the total population.

Although our simulation-based approach allowed us to robustly estimate key epidemiological parameters, we did not model other important aspects of university-community metapopulations affecting transmission dynamics. For example, changes in human mobility can change disease prevalence and lead to changes in behavioral patterns and contact rates [7]. Arrival testing was not mandated for WSU students returning to Whitman County and therefore the case importation rate was unknown. Knowing imported cases would have allowed for a more refined estimation of the time-varying reproductive number, which is very sensitive to imported cases and could have accounted for the slight differences between the ℛ_*t*_*estimates* and our Bayesian analyses. In addition, mobility patterns of community members in and out of Whitman Country were also not documented. Because this information was limited, we did not include migration in our mechanistic model. Instead, we began our analyses the week before the fall semester commenced when most students arrive and we could assume that the influx of students had reached an equilibrium. We also assumed movement in and out of the community population was minimal and at equilibrium. Another potential limitation of our approach was the inability to estimate all parameters in our model from the available Whitman County data. In addition to the transmission parameters that were estimated, our model also included parameters such as latency, recovery and positive testing rates and initial starting values. We did not estimate these parameters, but instead used values that were estimated independently from other studies. As such these parameters were estimated from data collected outside of Whitman County which could lead to bias in our results if Whitman County disease dynamics were largely dissimilar to other populations in Washington State and the U.S during the pandemic. Even though or mechanistic model may oversimplify reality and relies on results from data outside Whitman County, we balanced model tractability and parameter identifiablity with reasonable assumptions to estimate parameters and draw conclusions about transmission dynamics in a university town during fall 2020 of the COVID-19 pandemic.

Opinions converged during the second wave of COVID-19 in fall 2020 that the re-opening of many colleges and universities, which concentrated young adults in university towns, were to blame for high COVID-19 incidence [e.g., 13]. Outbreaks among university student and public concern even led to some institutions returning to online learning [11]. Retrospectively, our analyses support the hypothesis that systematic interventions, such as social distancing and masking, were highly effective in limiting SARS-CoV-2 transmission within and out-of university student populations even in the presence of heightened student mobility. Despite high, and occasionally headline-making case counts, our study suggests that the large peak in student cases was a pseudo-epidemic brought on by students returning to a single municipality with widespread access to testing, rather than widespread student-to-student transmission. In real time, this would be impossible to distinguish absent of arrival testing. This, in turn, suggests that arrival testing should be considered a key component of understanding the epidemiology of students returning to campus. Further, other universities and colleges may wish to revisit their ‘lessons learned’ regarding their pandemic control policies, if sheer numbers of cases were the basis by which those policies were evaluated. Health policy makers faced, and will continue to face, the dilemma of balancing public health with other competing interest such a the economy [7] and education with finite resources. We can now understand that keeping institutions of higher education open with mitigation strategies in place will not put co-located communities at excessive risk of COVID-19. This information will help us manage public health regulations for higher education in future pandemics of respiratory disease.

## Data Availability

All data, Mathematica Notebook and `R` code can be found at https://github.com/erinclancey/Trans-Dynam-Whit-Co.

## Acknowledgments

ETL, MSM and EC were funded by contracts 75D30121P10551, 75D30120P07911 and 75D30122C15691 from the Centers for Disease Control and Prevention, as well as RAPID 2110109 from the National Science Foundation. ETL was also supported on R35GM147013 from the National Institutes of Health.

