## Supplemental Material for "Unexpected Transmission Dynamics in a University Town: Lessons from COVID-19"

### Appendix A: Two-population model, derivation of $\mathcal{R}_0$, and specifying the generation interval

#### A.1 Two-population epidemic model

To study the transmission dynamics that occurred in Whitman County during the COVID-19 outbreak of fall 2020, we used a two-population Susceptible-Exposed-Infected-Recovered (SEIR) ordinary differential equations (ODE) model framework. The two-population ODE model specific to the university student and Whitman County community populations is

$$\dot{S}_u = -S_u(\beta_u I_u + \beta_m I_c) \tag{A1a}$$
$$\dot{S}_c = -S_c(\beta_c I_c + \beta_m I_u) \tag{A1b}$$
$$\dot{E}_u = S_u(\beta_u I_u + \beta_m I_c) - \sigma E_u \tag{A1c}$$
$$\dot{E}_c = S_c(\beta_c I_c + \beta_m I_u) - \sigma E_c \tag{A1d}$$
$$\dot{I}_u = \sigma E_u - \gamma I_u \tag{A1e}$$
$$\dot{I}_c = \sigma E_c - \gamma I_c \tag{A1f}$$
$$\dot{R}_u = \gamma I_u \tag{A1g}$$
$$\dot{R}_c = \gamma I_c. \tag{A1h}$$

We assumed the latency rate ($\sigma$) and recovery rate ($\gamma$) were equivalent in both populations, cross-transmission, $\beta_m$, from university students to community member is equal to community members to university students. We also assumed that population sizes remained constant throughout the duration of the fall semester, with no substantial loss to death or migration once the students arrived back on campus. Definitions of parameters and their symbols are given in Table 1 in the main text.

#### A.2 Derivation of $\mathcal{R}_0$

We derived $\mathcal{R}_0$ using the next-generation matrix (NGM), $\mathbf{K} = -\mathbf{T}\mathbf{\Sigma}^{-1}$, following [1]. $\mathcal{R}_0$ is the dominant eigenvalue of $-\mathbf{T}\mathbf{\Sigma}^{-1}$. $\mathbf{T}$ describes the transmission of new infections and $\mathbf{\Sigma}$ describes the transition out of the infection compartment. From the linearized infection subsystem in Equations (A1a,b,e,f) evaluated a the disease free equilibrium, we obtain

$$\mathbf{T} = \begin{pmatrix} S_{u_0}\beta_u & S_{u_0}\beta_m \\ S_{c_0}\beta_m & S_{c_0}\beta_j \end{pmatrix} \tag{A2}$$

and

$$\boldsymbol{\Sigma} = \begin{pmatrix} -\gamma & 0 \\ 0 & -\gamma \end{pmatrix}. \tag{A3}$$

Hence, the NGM $\mathbf{K}$ is given as

$$-\mathbf{T}\boldsymbol{\Sigma}^{\mathbf{-1}} = \begin{pmatrix} \frac{S_{u_0}\beta_u}{\gamma} & \frac{S_{u_0}\beta_m}{\gamma} \\ \frac{S_{c_0}\beta_m}{\gamma} & \frac{S_{c_0}\beta_j}{\gamma} \end{pmatrix}, \tag{A4}$$

where the dominant eigenvalue of $K$ gives us $\mathcal{R}_0$ in Equation (1) in the main text. All mathematical analyses were performed in Wolfram Mathematica 13.1 [2].

#### A.3 Specifying the generation interval distribution

To implement the methods in the package EpiEstim to estimate the time-varying reproductive number (Figure 2 in the main text), the user must supply the mean and standard deviation of the generation interval [3]. Following the mathematical framework presented in [4] and Clancey and Lofgren, the intrinsic generation interval for the two-population model in Equations (A1a-h) is

$$g(\tau) = \frac{\sigma\gamma}{\sigma - \gamma}\left(e^{-\gamma\tau} - e^{-\sigma\tau}\right), \tag{A5}$$

with mean

$$\mathbb{E}[\tau] = \frac{1}{\sigma} + \frac{1}{\gamma}, \tag{A6}$$

and variance

$$\mathbb{V}[\tau] = \frac{1}{\sigma^2} + \frac{1}{\gamma^2}. \tag{A7}$$

The numeric values for the mean and standard deviation of the generation interval used to produce Figure 2 in the main text were obtained using the parameter values for $\sigma$ and $\gamma$ in Table 1 in the main text.

### Appendix B: Bayesian Inference

#### B.1 Specifying the likelihood function

The series of ODEs in Equations A1a-h models the epidemic process in the overall population. To build a likelihood function for Bayesian inference, we must also model the sampling process that generated the observed COVID-19 case reports. First, we used an accumulator variable, $H$, to sum the simulated population incidence at each discrete time point 0 to $n$. Then, we sampled each $H_n$ to generate case reports at each time point using a negative binomial model,

$$\text{Case Reports}_n \sim \text{NegBinom}\Big(\rho H_n, k\Big). \tag{B1}$$

Here $\rho$ represents the portion of positive COVID-19 cases that were diagnosed and reported from the total number of case at each time point, $H_n$. We used the mean parameterization of the negative binomial distribution, with mean= $\rho H_n$ and variance= $\frac{\rho H_n + (\rho H_n)^2}{k}$, where $k$ is the overdispersion parameter. Equation (B1) forms the likelihood function conditioned on the epidemic process model in Equations (A1a-h)allowing us to sample from the posterior distributions for $\beta_u$, $\beta_c$, $\beta_m$, $\sigma$ and $\gamma$.

#### B.2 Simulation-based inference and posterior processing

We implemented the simulation-based estimation methods available in the `pomp` software package in R [5, 6] to generate maximum *a posteriori* (MAP) estimates and 95% highest posterior density interval (HPD) intervals for the unknown parameters (Table 2 in the main text). We used particle filtering MCMC (pMCMC) with 2,000 particles and a flat prior for each unknown parameter. We performed two sequential implementations of the estimation procedure to update the covariance matrix in the proposal distribution. In the first implementation we used independent Normal proposal distributions for each parameter. Using the posterior distribution from the first implementation, we estimated the covariance matrix between all unknown parameters. For all runs we initiated five chains, drawing the starting parameter values randomly, and ran each chain for 10,000 iterations. We used the Raftery and Lewis diagnostic [7] to determine the number of iterations to "burn-in" (all unknown parameters had a suggested burn-in<50). We assessed within chain convergence using the Geweke diagnostic [8] and global convergence using the the Gelman diagnostic [9]. All chains converged successfully within and across chains. After the second implementation of the estimation procedure, we discarded the first 50 iterations as "burn-in" and thinned each chain over a 50 sample window, leaving us with 1,000 samples in the joint posterior distribution. Processing of the posterior distributions was performed using the package *coda* in R [10].

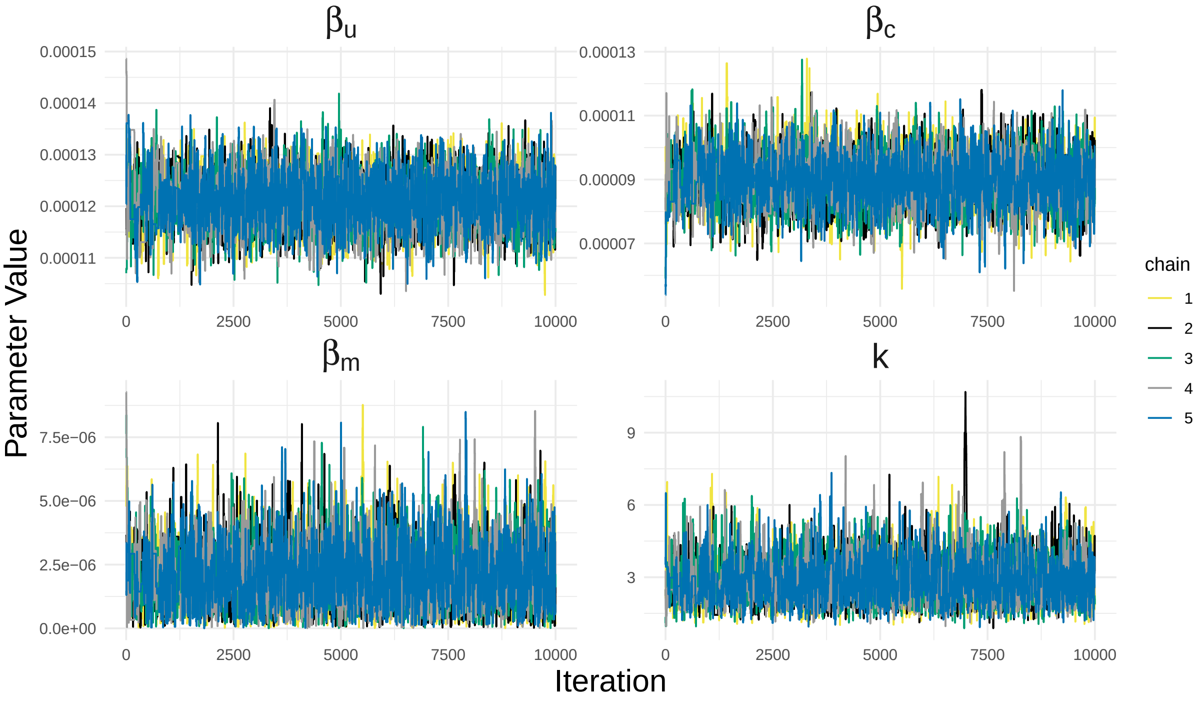


Figure B1: Trace plot of the five chains over 10,000 iterations for each parameter estimated in the model.

We estimated point (MAP) and intervals (HDI) for each unknown parameter in our model. MAP's were estimated as the marginal posterior modes using kernel density estimation with a bandwidth of 1 from the package *MCMCglmm: MCMC Generalised Linear Mixed Models* in R [11]. We obtained the 95% HDIs in R using *bayestestR* [12]. Numeric values of these estimates are given in Table 2 in the main text.

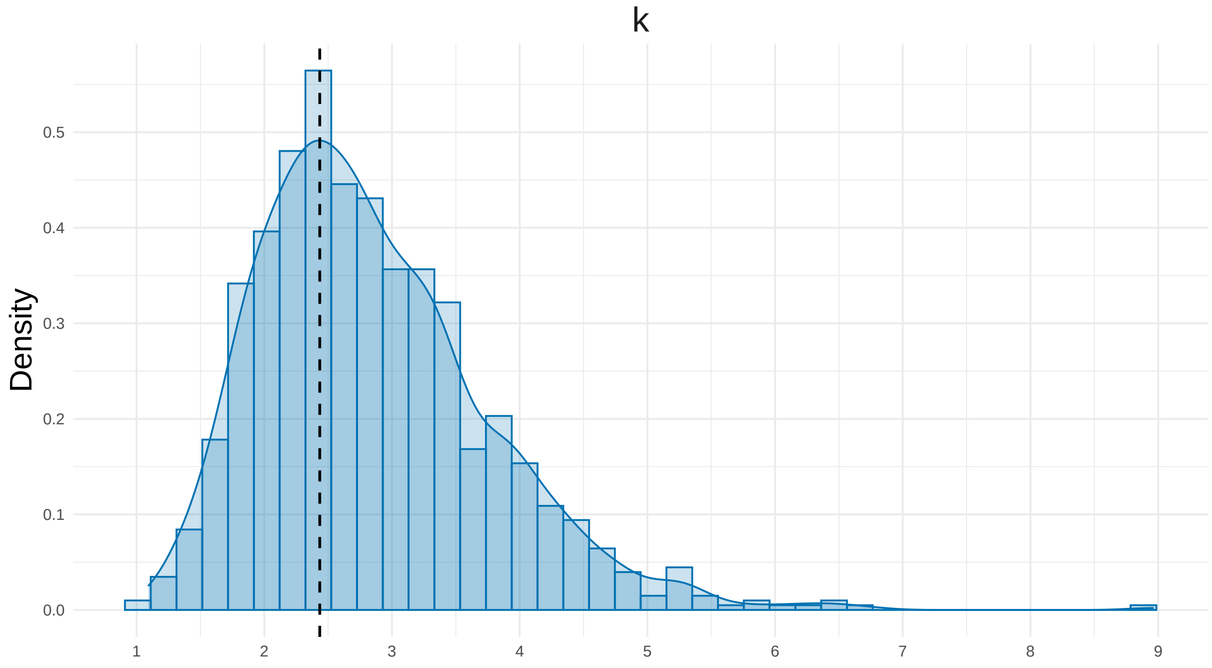


Figure B2: Marginal posterior density for $k$. The black dashed line represents the posterior mode.

[2] Wolfram Research, Inc. Mathematica, 2021. Place: Champaign, Illinois.

[3] Anne Cori, Neil M. Ferguson, Christophe Fraser, and Simon Cauchemez. A new framework and software to estimate time-varying reproduction numbers during epidemics. *American journal of epidemiology*, 178(9):1505–1512, 2013. Publisher: Oxford University Press.

[4] David Champredon, Jonathan Dushoff, and David J. D. Earn. Equivalence of the Erlang-Distributed SEIR Epidemic Model and the Renewal Equation. *SIAM Journal on Applied Mathematics*, 78(6):3258–3278, January 2018.

[5] Aaron A. King, Dao Nguyen, and Edward L. Ionides. Statistical Inference for Partially Observed Markov Processes via the R Package pomp. *Journal of Statistical Software*, 69(12), 2016.

[6] R Core Team. R: A Language and Environment for Statistical Computing, 2023.

[7] AL Raferty and S. Lewis. One long run with diagnostics: implementation strategies for Markov chain Monte Carlo. *Statistical Science*, 7(4):475.
